# Reliable Uncertainty Under Class Imbalance and Distribution Shift: Class-Conditional Conformal Prediction of Multiple Sclerosis

**DOI:** 10.64898/2026.05.12.26353057

**Authors:** Alexander S. Millar, Cortnee Román, Ramkiran Gouripeddi, Julio C. Facelli

## Abstract

**Objectives:** To evaluate whether class-conditional conformal prediction (CP) can provide reliable uncertainty quantification (UQ) under severe class imbalance and distribution shift, using multiple sclerosis (MS) diagnosis from magnetic resonance imaging (MRI) as a clinical exemplar.

**Methods:** We evaluated marginal and class-conditional CP using 720 T2-weighted MRI scans (142 MS, 578 controls). A convolutional neural network trained on 3 T data was evaluated under distribution shift (1.5 T acquisitions and synthetic image degradations). Through 100 Monte Carlo experiments, we assessed coverage guarantees, class-specific performance, and relationships between calibration set size, coverage variance, and uncertainty.

**Results:** Marginal CP severely under-covered the minority MS class (16.9% mean coverage at 1.5 T vs. 95.2% for controls) despite valid population-level guarantees. Class-conditional CP dramatically improved MS coverage to 77.5% at 1.5 T and 85.8% at 3 T, significantly reducing severe undercoverage (<80%) frequency while maintaining >89% control coverage. Minority class coverage variance increased due to limited calibration samples, matching theoretical Beta-binomial predictions. CP maintained validity under distribution shift; prediction set sizes scaled monotonically with shift severity, yielding clinically interpretable UQ.

**Conclusions:** Class-conditional CP successfully mitigates systematic undercoverage of minority disease classes while maintaining validity under distribution shift. The approach offers a practical, model-agnostic solution for uncertainty quantification applicable across clinical AI systems, though increased coverage variance for less represented conditions reflects fundamental statistical constraints. By characterizing these variance trade-offs, this framework enables more reliable deployment of diagnostic AI in heterogeneous clinical environments across diverse medical domains where minority disease class detection is critical.

## 1 Introduction

Artificial intelligence (AI) systems for medical image analysis have demonstrated remarkable diagnostic accuracy across numerous conditions, yet their clinical deployment remains limited by fundamental challenges in uncertainty quantification (UQ) at the level of individual patients [1], [2]. While clinical AI models can match or exceed overall specialist performance in controlled settings, they often fail when deployed in real-world environments characterized by heterogeneous patient populations, varying imaging protocols, and evolving disease presentations [3], [4], [5], [6], [7], [8] Translational efforts to bridge the development-to-deployment gap are particularly challenged in low-prevalence and underrepresented diseases, where severe class imbalance and limited training examples create compounding UQ challenges that standard machine learning approaches cannot adequately address [9].

Multiple sclerosis (MS) exemplifies these challenges. This chronic neurological condition affects approximately 2.8 million people worldwide, with prevalence varying from 5 to 300 per 100,000 depending on geographic location [10], [11], [12]. Early diagnosis critically impacts patient outcomes, as disease-modifying therapies are most effective when initiated before significant neurological damage accumulates [13], [14], [15]. Magnetic Resonance Imaging (MRI) is central to MS diagnosis, with lesions detectable in the brain, spinal cord, and optic nerves. Brain MRI remains the primary diagnostic tool, revealing characteristic lesions in periventricular, juxtacortical, and infratentorial white matter regions [16], [17]. Yet the subtle, variable nature of early MS lesions, combined with the relative rarity of the condition in screening populations, creates substantial diagnostic difficulty. These challenges are magnified when AI systems trained on high-quality academic datasets must generalize to diverse clinical environments with varying field strengths, acquisition protocols, and image quality standards [7], [18].

The biomedical domain has not established clear UQ frameworks despite ubiquitous data uncertainties. Fewer than 4% of medical AI studies explicitly address uncertainty in their predictions at the patient level [19], [20], reflecting a critical gap between the recognized importance of patient-level uncertainty quantification and its actual implementation. When AI systems systematically underestimate uncertainty for minority disease classes, they create a dangerous illusion of confidence precisely where clinical vigilance is most needed [9]. For MS, a false negative means delayed diagnosis and missed opportunities for early intervention, potentially resulting in irreversible neurological damage, disability accumulation, and increasing healthcare costs due to progressive disability. Conversely, false positives trigger unnecessary anxiety, expensive follow-up testing, and potential exposure to inappropriate treatments. Without reliable uncertainty estimates that maintain validity across diverse patient subpopulations—particularly those underrepresented in training data—clinicians cannot appropriately calibrate their trust in AI recommendations [21].

A central barrier to clinical AI translation is distribution shift: MRI-based models trained on high-quality academic medical center imaging must often operate on lower-quality community hospital acquisitions that differ in scanner type, protocol, or patient population. For MS diagnosis, such shifts are especially pronounced—the move from 3 T to 1.5 T scanners reduces contrast and signal-to-noise, while motion, field inhomogeneities, and post-processing add blur and variability [22]. Machine learning’s assumption that training and test data share the same distribution is violated under these conditions, causing performance metrics validated on held-out test sets to misrepresent deployment reliability [7], [23].

Many existing UQ methods prove impractical for clinical deployment. Bayesian approaches require adopting specific probabilistic architectures and retraining from scratch, while ensemble methods necessitate training and maintaining multiple models [24], [25], [26], [27], [28]. Techniques relying on access to training distributions or model internals are incompatible with the black-box, proprietary AI systems already in widespread use [29], [30], [31]. The clinical community therefore requires practically implementable frameworks that move beyond average performance metrics to assess reliability at the individual patient level.

Among various approaches, conformal prediction (CP) emerges as particularly suited to clinical deployment [21]. CP is a model-agnostic statistical framework providing distribution-free theoretical guarantees—reliable predictions regardless of underlying data distribution, assuming exchangeability [32], [33]. Unlike traditional confidence intervals relying on rarely-met distributional assumptions, CP constructs prediction sets that contain the true outcome with a specified probability [34], [35]. This post-hoc method can be layered onto any predictive system without model modifications, making it suitable for third-party medical device software where model access may be limited [34].

However, CP’s guarantees are typically marginal, averaging across the entire population. In an imbalanced clinical dataset, a system may achieve nominal 90% coverage by providing near-perfect coverage for abundant healthy patients while systematically failing for the minority disease class [36]. To address this, class-conditional conformal prediction (Mondrian CP) calibrates error rates separately for each class, ensuring coverage guarantees hold within each subgroup rather than only in aggregate [32], [35], [37], [38]. This approach has shown promise in stabilizing uncertainty quantification in imbalanced datasets [38], [39].

Recent work by Sreenivasan et al. applied Mondrian CP to distinguish between MS subtypes using electronic health record data, demonstrating individualized diagnostic uncertainty [40]. However, their study addressed classification within an MS-only cohort and did not confront the severe imbalances inherent in disease-vs-healthy screening tasks. Significant challenges remain in extending these methods to the highly skewed settings characteristic of low-prevalence disease detection under distribution shift.

### 1.1 Study Objectives and Hypothesis

To address this gap, this study investigates whether class-conditional CP can provide reliable UQ for MS diagnosis from MRI data under the challenging conditions typical of clinical deployment: severe class imbalance and distribution shift between training and deployment environments. Our primary objective is to determine whether class-conditional methods can mitigate the coverage failures of standard CP for the minority class (MS patients), while maintaining valid uncertainty estimates as deployment conditions diverge from training. We hypothesize that class-conditional CP will (1) substantially reduce coverage imbalance between the data from healthy controls and MS patients compared to marginal methods, (2) maintain these improved coverage properties under distribution shift from 3 T training data to 1.5 T deployment data and various image quality degradations, and (3) reveal interpretable relationships between calibration set size, coverage variance, and prediction set size that inform clinical deployment strategies. Through systematic evaluation across 100 Monte Carlo experiments with multiple distribution shift scenarios, we aim to establish whether CP methods can provide the robust uncertainty quantification necessary for the safe deployment of AI diagnostic systems in heterogeneous clinical environments.

## 2 Methods

### 2.1 Study Design and Datasets

We evaluated CP methods for UQ in MS diagnosis using T2-weighted MRI data from multiple sources. The study utilized 720 scans: 142 scans of MS patients and 578 healthy controls across different field strengths and acquisition sites. Training data comprised 61 scans from the (International Symposium on Biomedical Imaging) ISBI 2015 MS Lesion Segmentation Challenge test subset (3 T, Philips Medical Systems) and 61 randomly selected healthy controls from the (Information eXtraction from Images) IXI dataset’s Hammersmith Hospital site (3 T, Philips Intera) [41], [42]. The remaining 567 scans served as the evaluation set, including 21 MS scans from ISBI training data, 29 MS scans from the Muslim et al. Iraqi cohort (1.5 T, multiple sites), and 548 healthy controls from IXI (124 from Hammersmith 3 T, 319 from Guy’s Hospital 1.5 T Philips, 74 from Institute of Psychiatry 1.5 T GE) [43]. Ground truth diagnoses were established by each dataset’s original clinical criteria.

### 2.2 Image Preprocessing and Standardization

All T2-weighted scans underwent standardized preprocessing using Advanced Normalization Tools (ANTs v2.5.4) [44]. Initial corrections addressed orientation inconsistencies in the Muslim et al. dataset, where 60 scans required 90-degree axial rotation and voxel dimension corrections based on [43] provided supplementary acquisition parameters. Following orientation correction, all scans underwent N4 bias field correction with parameters optimized for T2-weighted imaging (shrink factor=2, convergence=[50×50×50×50, 0.0001], B-spline distance=200) [45].

Rigid registration to the MNI152NLin2009cAsym T2-weighted template (1mm isotropic) employed mutual information as the similarity metric, chosen for robustness across multi-site data [46], [47]. Registration parameters included multi-resolution optimization (12×8×4×2 shrink factors), intensity winsorization [0.005, 0.995], and histogram matching. Quality control excluded 31 Muslim et al. scans due to unacceptable post-registration orientation or tissue loss.

For each registered volume, we selected 43 contiguous axial slices capturing anatomical regions relevant to MS diagnosis: lateral ventricles, periventricular white matter, corpus callosum, and juxtacortical regions. Slice selection began immediately inferior to the characteristic "butterfly" appearance of the lateral ventricles. Selected slices were exported as 8-bit grayscale images for subsequent processing.

### 2.3 Distribution Shift Simulation

To evaluate robustness under realistic deployment scenarios, we created seven synthetic variants of each scan following established protocols [22]. Progressive Gaussian blur with standard deviations (𝜎) of 1, 2, 3, and 4 pixels simulated motion artifacts and resolution degradation typical of patient movement or lower-quality acquisitions. Three contrast modifications represented common post-processing variations: adaptive histogram equalization (CLAHE) for local contrast enhancement, global histogram equalization for intensity redistribution, and intensity adjustment (imadjust) using linear contrast stretching. These transformations (Figure 1), implemented in MATLAB R2023b, created 16 distinct evaluation conditions when combined with the two field strengths (8 variants × 2 field strengths).

**Figure 1.**
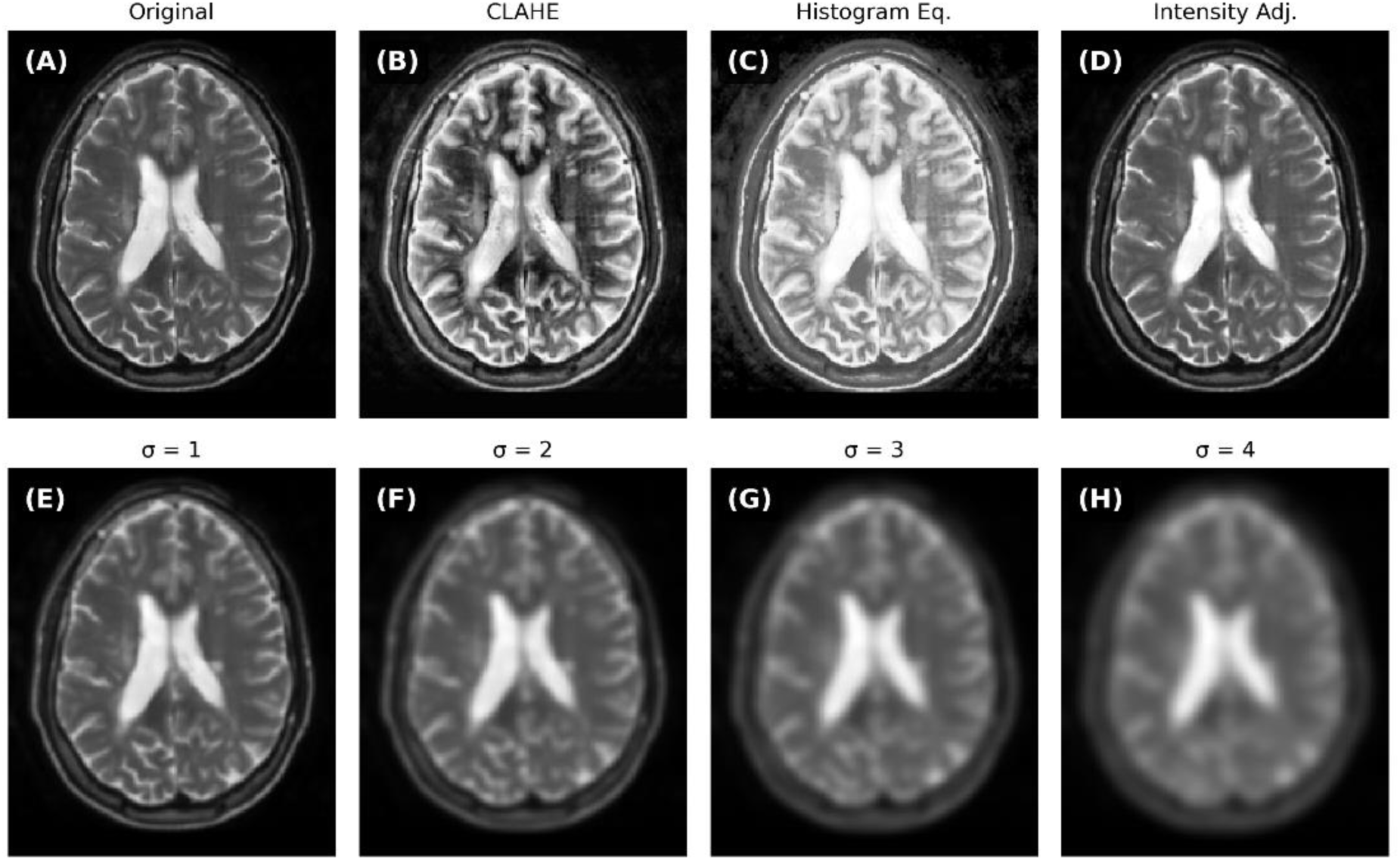
Representative examples of distribution shift transformations applied to T2-weighted MRI data. Top row: Contrast modifications simulating post-processing variations. (**A**) Original baseline, (**B**) Adaptive histogram equalization (CLAHE), (**C**) Global histogram equalization, (**D**) Intensity adjustment (imadjust) with 1%-99% saturation limits. Bottom row: Progressive Gaussian blur simulating motion artifacts and resolution degradation. (**E-H**) Gaussian blur with 𝜎 = 1, 2, 3, and 4 pixels respectively, showing increasing loss of anatomical detail and tissue boundary definition.

### 2.4 Deep Learning Model Architecture

We employed a fully convolutional network architecture previously validated for MS classification. The network consisted of five convolutional blocks with progressively increasing filter counts (50, 256, 512, 1024, 2), using kernel sizes of 11×11, 5×5, 3×3, 2×2, and 1×1 respectively [22]. Batch normalization followed each convolutional layer except the final classification head. Dropout (rate=0.5) was applied before the spatially-resolved classification layer to reduce overfitting. Global max pooling aggregated spatial features for slice-level predictions. The model was implemented in TensorFlow 2.18.0 and trained using Adam optimization (learning rate=1e-3 with cosine decay), sparse categorical cross-entropy loss, and early stopping (patience=10 epochs).

### 2.5 Training Protocol

Training employed extensive data augmentation to prevent overfitting and improve generalization. The augmentation pipeline included random brightness/contrast adjustment (limit = 0.2), gamma correction (80-120), CLAHE (clip_limit = 2.0), Gaussian blur (blur_limit = (3, 5)), JPEG compression (quality = 50-80), Gaussian noise (variance = 0.0005-0.002), and mild geometric transformations (shift ≤ 2%, scale ≤ 3%, rotation ≤ 5°). Following augmentation, all images were center-cropped to 192×192 pixels and normalized to [0,1] intensity range. Training proceeded for up to 50 epochs with batch size 32, using model checkpointing to preserve best validation performance.

### 2.6 Conformal Prediction Implementation

We implemented both marginal and class-conditional split-conformal prediction following established frameworks. Nonconformity scores were computed as 𝛼_𝑖_ = 1 − 𝑓(𝑥_𝑖_)_𝑦𝑖_, where 𝑓(𝑥_𝑖_)𝑦_𝑖_ represents the model’s softmax probability for the true class. For each test example and candidate class 𝑦, conformal p-values were calculated as the proportion of calibration nonconformity scores greater than or equal to 1 −𝑓(𝑥)_𝑦_, with finite-sample correction applied by adding 1 to both numerator and denominator. Prediction sets at significance level 𝛼 = 0.1 (corresponding to 90% target coverage) included all classes 𝑦 with p-values exceeding 𝛼. Credibility was defined as the conformal p-value of the most likely candidate class, and confidence as one minus the p-value of the next-best alternative. Class-conditional CP maintained separate nonconformity score distributions per class, computing p-values using only calibration examples from the corresponding class, ensuring coverage guarantees are held conditionally within each class rather than only marginally across the population.

### 2.7 Experimental Design

We conducted 100 Monte Carlo experiments with random calibration-test splits of the evaluation data, following established recommendations for evaluating CP implementations that suggest this number of iterations to properly account for finite-sample variability and enable reliable assessments [35]. Each iteration randomly selected 42 scans (1,806 slices) for calibration, with algorithmic verification ensuring both classes were represented to prevent degenerate prediction sets. The remaining scans formed the test set.

Our analysis focused on three key aspects of conformal prediction performance. First, we evaluated whether marginal coverage guarantees were maintained as deployment conditions diverged from training, examining performance across eight data variants (baseline plus seven synthetic transformations) at both 3 T and 1.5 T field strengths. Second, we assessed coverage balance between classes, comparing marginal and class-conditional methods on baseline data to quantify coverage disparities for the minority MS class. Third, we investigated the relationship between calibration set size and coverage variance, using theoretical finite-sample distributions to validate observed patterns [35]. This design enabled evaluation of both statistical validity and clinical utility under realistic deployment scenarios.

### 2.8 Evaluation Metrics

Coverage analysis assessed three metrics: empirical coverage (proportion of correct predictions), undercoverage frequency (coverage <90%), and severe undercoverage frequency (coverage <80%). Prediction set sizes quantified uncertainty magnitude and clinical utility. We computed theoretical finite-sample coverage distributions using Beta-binomial models parameterized by calibration and test set sizes to validate observed variance patterns. Additional metrics included confidence (1 minus second-highest conformal p-value), credibility (highest conformal p-value), and margin (difference between top two conformal p-values).

### 2.9 Statistical Analysis and Software

All analyses employed Python 3.9 with custom implementations based on established conformal prediction frameworks. Statistical comparisons used median values across Monte Carlo runs to ensure robustness to outliers. Coverage guarantees were evaluated at both population (marginal) and class-specific levels. Results report median coverage and interquartile ranges across experimental runs. Computational experiments utilized resources from the Center for High Performance Computing at the University of Utah. Complete code, the trained model, and full documentation are available at https://github.com/illato/mricp.

## 3 Results

We first evaluated whether standard (marginal) CP could provide reliable UQ for MS diagnosis from MRI data. Using 100 independent calibration-test splits of baseline data, we found that marginal CP failed for the minority MS class while over-covering healthy controls (Figure 2, Table 1). At 1.5 T field strength, MS patients achieved only 16.9% mean coverage compared to 95.2% for controls—a coverage gap of 78.3 percentage points. Even at 3 T, where class imbalance was less severe, MS patients received only 54.2% coverage versus 94.6% for controls. Critically, 100% of experimental runs resulted in severe undercoverage (<80%) for MS patients at 1.5 T, and 99% at 3 T, representing complete failure of uncertainty quantification for the clinically relevant minority class.

**Figure 2.**
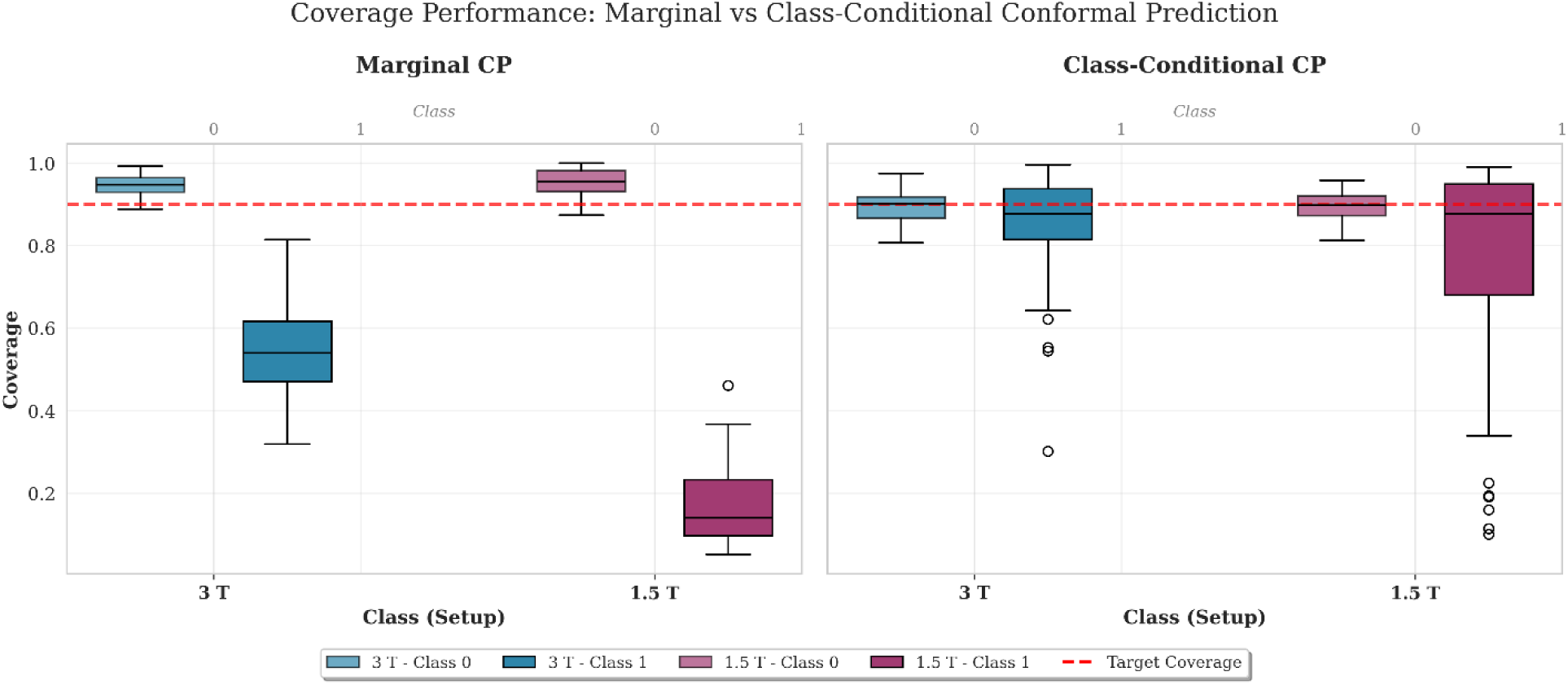
Coverage performance comparison between marginal and class-conditional CP across field strengths and diagnostic classes. Box plots show the distribution of coverage values across 100 calibration/test splits for each experimental condition for the unmodified baseline data. The red dashed line indicates the target coverage of 0.90. Left panel shows marginal CP (NC), right panel shows class-conditional CP (CC). Class 0 represents control subjects; Class 1 represents MS patients. Field strengths: 3 T (blue) and 1.5 T (purple), with transparency distinguishing between classes (lighter for controls, darker for MS patients).

**Table 1.** Quantitative comparison of coverage statistics between marginal (NC) and class-conditional (CC) conformal prediction methods.

| Field Strength | Type | Class | Mean Coverage | Median Coverage | Coverage SD | Undercoverage (< 0.90) | Severe Undercoverage (< 0.80) |
| --- | --- | --- | --- | --- | --- | --- | --- |
| 1.5 T | NC | 0 | 0.952 | 0.954 | 0.032 | 6% | 0% |
|  |  | 1 | 0.169 | 0.142 | 0.087 | 100% | 100% |
|  | CC | 0 | 0.895 | 0.898 | 0.033 | 51% | 0% |
|  |  | 1 | 0.775 | 0.878 | 0.240 | 56% | 37% |
| 3.0 T | NC | 0 | 0.946 | 0.948 | 0.024 | 3% | 0% |
|  |  | 1 | 0.542 | 0.540 | 0.112 | 100% | 99% |
|  | CC | 0 | 0.894 | 0.901 | 0.036 | 49% | 0% |
|  |  | 1 | 0.858 | 0.877 | 0.113 | 59% | 22% |

*Statistics computed across 100 independent calibration/test splits of unmodified baseline data across two field strengths. Undercoverage defined as coverage < 0.90; severe undercoverage as coverage < 0.80. Field strengths: 1.5 T and 3.0T. Class 0 = controls, Class 1 = MS patients.*

Class-conditional CP dramatically improved coverage equity between classes. For MS patients at 1.5 T, mean coverage increased from 16.9% to 77.5%, while at 3 T it improved from 54.2% to 85.8%. The frequency of severe undercoverage for MS patients dropped from 100% to 37% at 1.5 T and from 99% to 22% at 3 T (Figure 2). This improvement came at a modest cost to the majority class: controls maintained mean coverage above 89% at both field strengths, with no instances of severe undercoverage. The increased undercoverage frequency for controls (from 6% to 51% at 1.5 T) primarily reflected minor deviations below the 90% target rather than clinically significant coverage failures.

The improved coverage for MS patients under class-conditional methods came with substantially increased variance (Figure 3). For the minority MS class, coverage distributions showed interquartile ranges spanning 15-25 percentage points at 1.5 T and 10-15 percentage points at 3 T, compared to tight distributions for healthy controls. This variance directly correlated with the effective calibration set sizes: MS patients had only 123 calibration slices at 1.5 T versus 1,682 for controls—a 13.7-fold difference (Table 2). The 3 T dataset, with its 6.1-fold difference (253 vs 1,552 slices), showed correspondingly reduced but still substantial variance.

**Figure 3.**
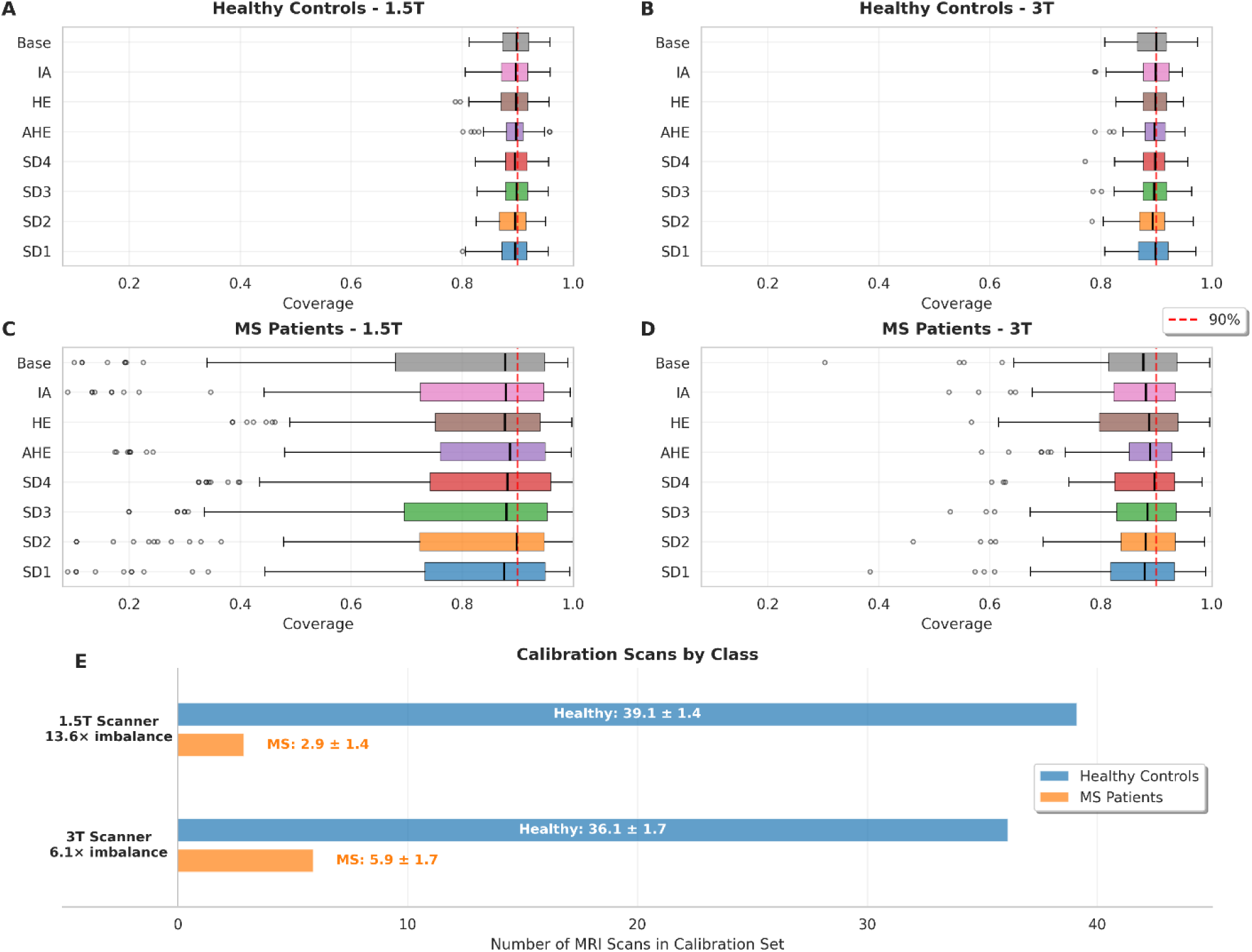
Class-conditional CP coverage varies inversely with calibration set size. Empirical coverage distributions across 100 Monte Carlo experiments demonstrate the impact of class imbalance on prediction interval reliability. **(A, B)** Coverage for the majority class (healthy controls) remains tightly concentrated around the nominal 90% level (dashed red line) for both 1.5 T (A) and 3 T (B) field strengths across all data variants including baseline and systematically degraded images, including progressive Gaussian blur (SD1-4) and contrast perturbations (AHE: adaptive histogram equalization; HE: histogram equalization; IA: intensity adjustment). **(C, D)** Coverage for the minority class (MS patients) exhibits substantially greater variance, with wider interquartile ranges and more extreme outliers, particularly pronounced in the 1.5 T data (C) where the MS class comprises only 7% of calibration samples. **(E)** Distribution of calibration set sizes by class reveals the source of coverage variance. The 13.6-fold difference in calibration samples between classes in 1.5 T data drives the observed coverage instability, while the more balanced 6.1-fold difference in 3 T data results in comparatively reduced variance. Each box represents 100 independent calibration-test splits.

**Table 2.**
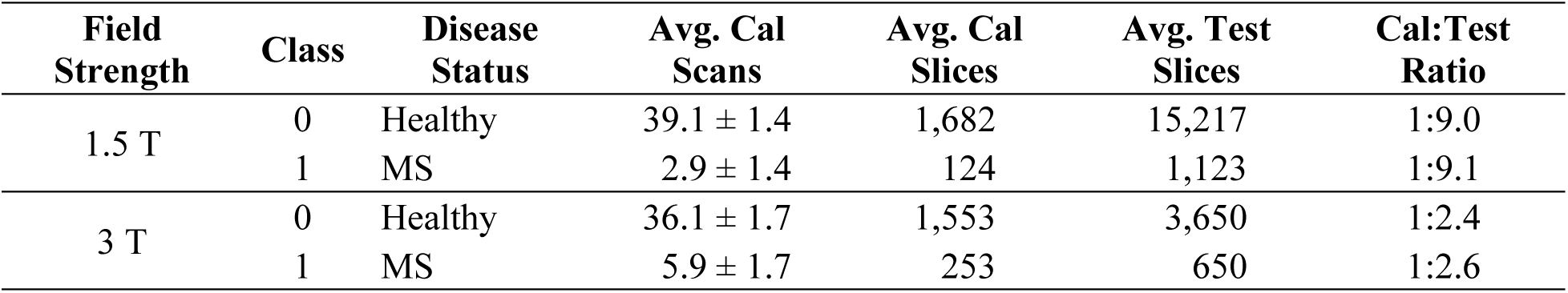
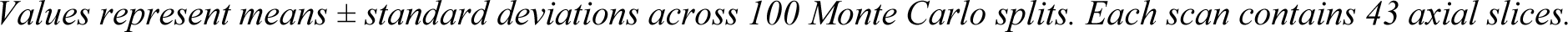
Calibration and test set composition by class and field strength.

| Field Strength | Class | Disease Status | Avg. Cal Scans | Avg. Cal Slices | Avg. Test Slices | Cal:Test Ratio |
| --- | --- | --- | --- | --- | --- | --- |
| 1.5 T | 0 | Healthy | $39.1 \pm 1.4$ | 1,682 | 15,217 | 1:9.0 |
| | 1 | MS | $2.9 \pm 1.4$ | 124 | 1,123 | 1:9.1 |
| 3 T | 0 | Healthy | $36.1 \pm 1.7$ | 1,553 | 3,650 | 1:2.4 |
| | 1 | MS | $5.9 \pm 1.7$ | 253 | 650 | 1:2.6 |

Values represent means ± standard deviations across 100 Monte Carlo splits. Each scan contains 43 axial slices.

Theoretical analysis confirmed this variance as an expected statistical property rather than a methodological failure (Figure 4). The Beta-binomial distributions governing finite-sample coverage closely matched our empirical observations, demonstrating that increased uncertainty for minority classes arises naturally from quantile estimation with limited samples. This fundamental constraint cannot be eliminated by methodological refinements alone—it reflects genuine statistical uncertainty when calibrating on small minority class samples.

**Figure 4.**
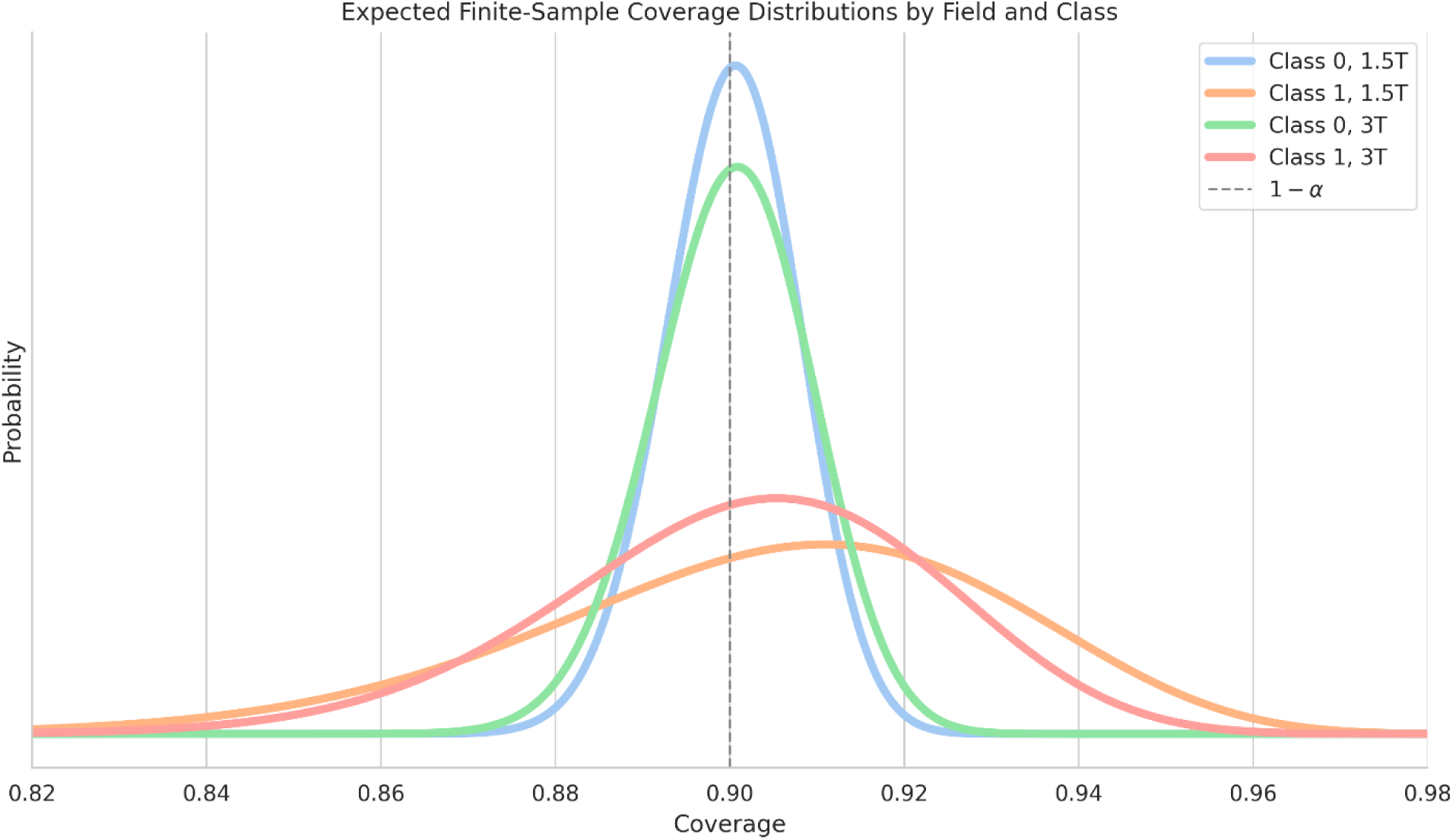
Theoretical finite-sample coverage distributions predict observed empirical variance. Beta-binomial distributions derived from average calibration and test set sizes demonstrate that coverage variance is an expected statistical property rather than a methodological limitation. Distributions are computed using parameters α = n + 1 -⌊(n + 1)α⌋ and β = ⌊(n + 1)α⌋, where n represents calibration set size and α = 0.1 is the pre-specified error rate. The minority MS class shows broader expected coverage distributions in both 1.5 T (orange) and 3 T (red) datasets compared to the majority healthy class (blue and green, respectively), with distribution width inversely proportional to calibration set size. Probability densities are normalized by the ratio of test set sizes to facilitate visual comparison. The vertical dashed line indicates the nominal coverage level (1 - α = 0.9). These theoretical distributions closely match the empirical variance observed in Figure 3, confirming that increased coverage uncertainty for minority classes is a fundamental finite-sample property of conformal prediction.

To assess robustness under realistic deployment scenarios, we evaluated both marginal and class-conditional methods across simulated distribution shifts representing common sources of clinical variation. Despite training exclusively on 3 T baseline MRI data, CP maintained valid marginal coverage across all conditions tested, including 1.5 T acquisitions and various image degradations (Figure 5).

**Figure 5.**
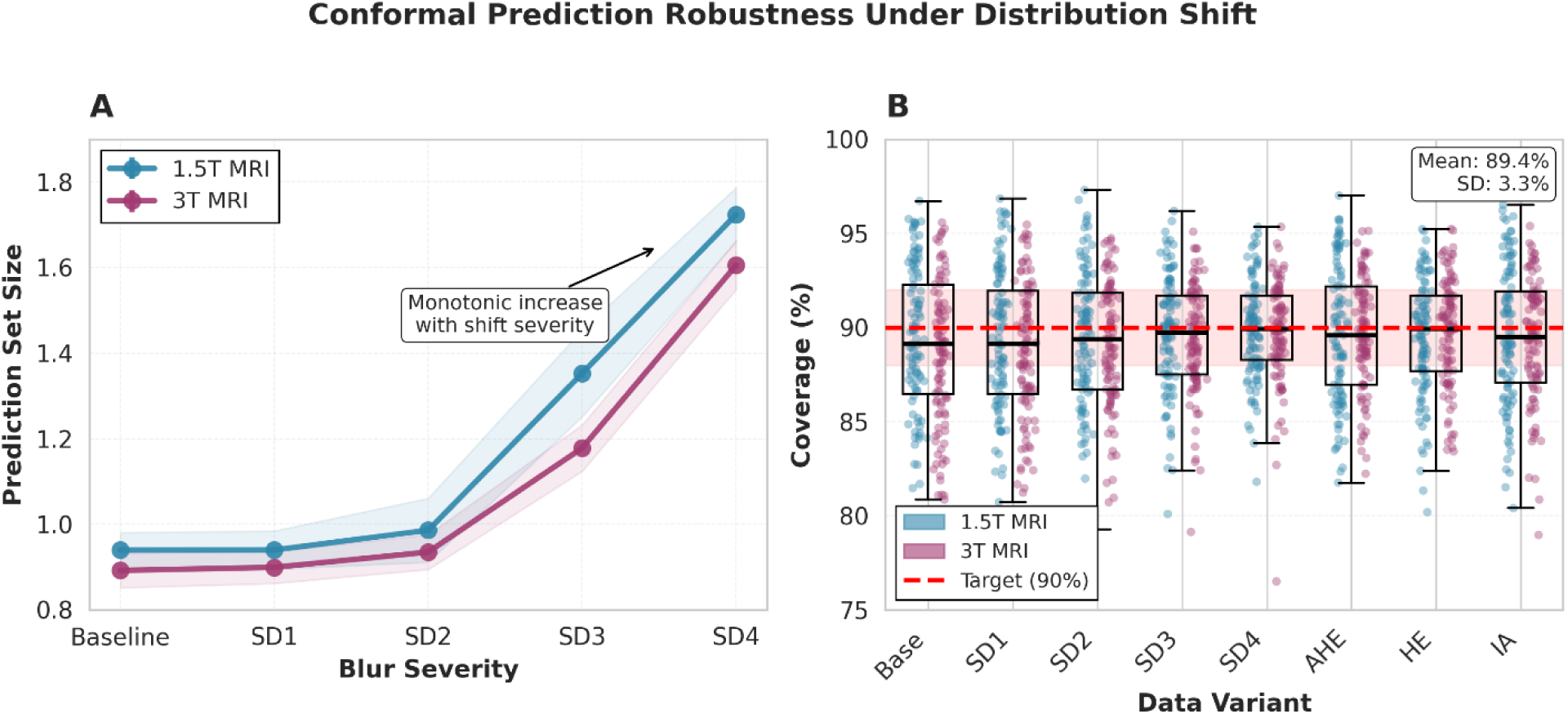
CP maintains statistical validity while appropriately increasing uncertainty under distribution shift. **(A)** Dose-response relationship between blur severity and prediction set size for 1.5 T and 3 T MRI data. Lines show mean values with standard error bands across 100 independent calibration-test splits. Prediction set size increases monotonically with distribution shift severity, expanding by 83.4% (1.5 T) and 79.9% (3 T) from baseline to maximum blur (SD4). **(B)** Empirical coverage across all distribution shift variants, demonstrating preservation of the 90% nominal coverage guarantee (red dashed line). Individual points represent single experimental runs, with box plots showing statistics across field strengths. Despite substantial variation in prediction set sizes across conditions, mean coverage remains stable at 89.6% ± 3.5% (1.5 T) and 89.3% ± 3.2% (3 T), validating the robustness of conformal guarantees under covariate shift. Data variants include progressive Gaussian blur (SD1-4) and contrast modifications (AHE: adaptive histogram equalization; HE: histogram equalization; IA: intensity adjustment).

The preservation of coverage guarantees came at the cost of increasingly conservative predictions as the deployment distribution diverged from training conditions (Supplementary Table S1). Prediction set sizes expanded monotonically with blur severity, increasing by 83.4% for 1.5 T and 79.9% for 3 T under maximum degradation (SD4). Remarkably, empirical coverage remained stable at 89.6% ± 3.5% (1.5 T) and 89.3% ± 3.2% (3 T) across all conditions, validating theoretical guarantees even under severe covariate shift.

The graduated response to distribution shift revealed an important clinical utility: prediction set size itself serves as a diagnostic metric for input quality. Mild degradations produced proportionally mild increases in uncertainty, while severe degradations appropriately triggered more conservative predictions. This predictable behavior aligns with clinical intuition that poorer image quality should reduce diagnostic confidence.

Analysis of individual predictions demonstrated how conformal methods translate statistical guarantees into clinically interpretable outputs through prediction sets and conformal uncertainty measures (Figure 6). Under marginal conformal prediction, uncertain cases often returned empty prediction sets ("I don’t know" [19], [25]), while class-conditional methods provided graduated uncertainty through variable set sizes.

**Figure 6.**
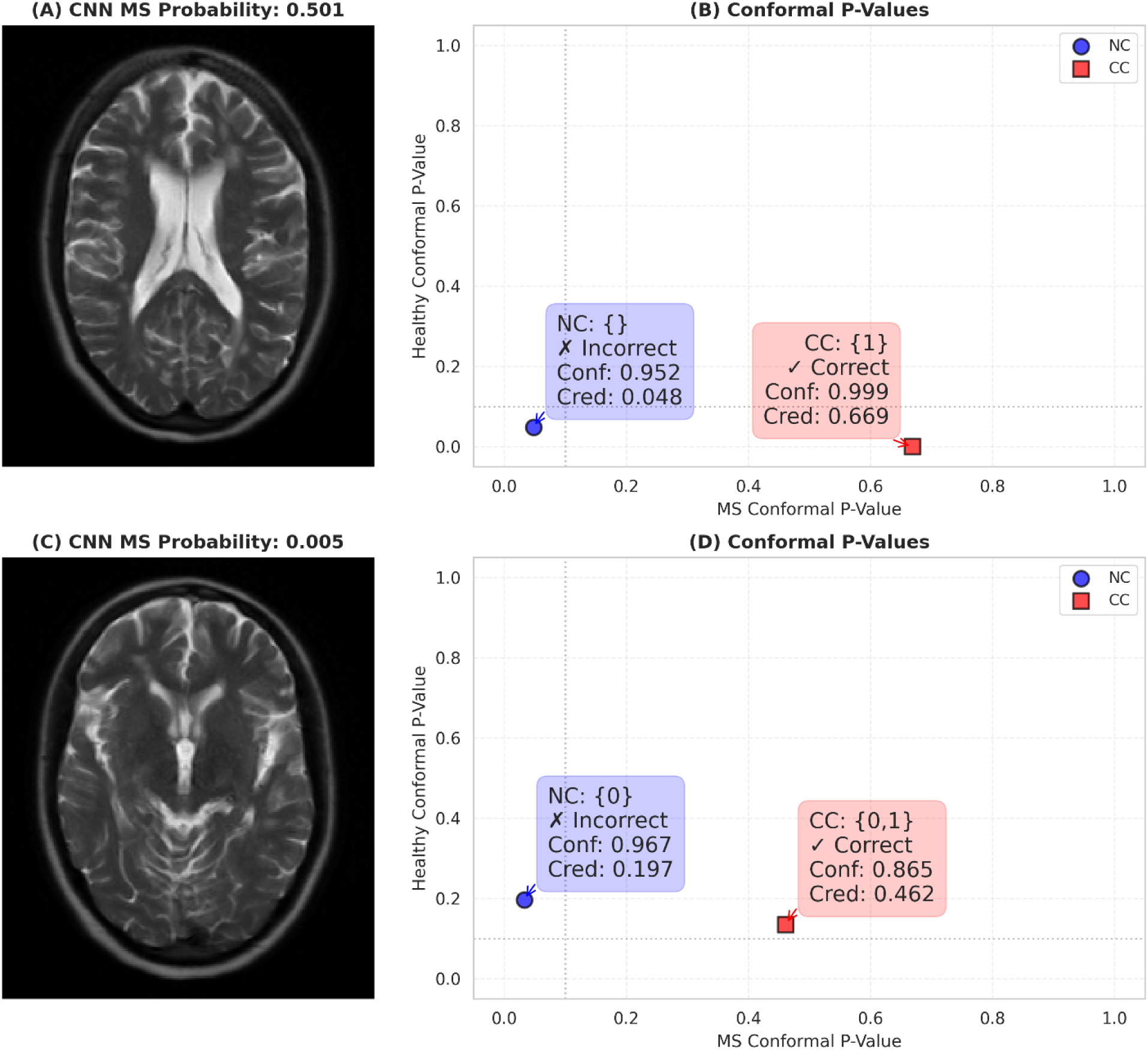
Case studies illustrating how class-conditional conformal prediction (CC) refines uncertainty compared with the marginal (NC) method. (**A**) T2-weighted MRI slice from a patient with MS, where the convolutional neural network (CNN) assigns an ambiguous probability of 0.501. (**B**) Conformal p-values at α = 0.1 (dotted lines) show that NC produces an empty prediction set {} (confidence = 0.952, credibility = 0.048), while CC yields a decisive singleton {1} (MS) with higher credibility (0.669). (**C**) T2-weighted MRI slice from a patient with MS, where the CNN incorrectly assigns a very low MS probability of 0.005, strongly favoring the wrong class. (**D**) Conformal p-values show that NC incorrectly predicts only the healthy class {0} (confidence = 0.967, credibility = 0.197), whereas CC includes both classes {0,1}, preserving valid coverage (confidence = 0.865, credibility = 0.462) and protecting against systematic undercoverage of the minority MS class when the model is overconfident but incorrect.

Singleton predictions—where the prediction set reduces to a single class—offered an especially intuitive lens. Confidence (1 – second-highest p-value) reflects whether plausible alternatives remain, while credibility (the maximum p-value) gauges how typical the case is relative to the calibration distribution. A high-confidence singleton, as in Figure 6b, indicates a decisive prediction; its moderate credibility signals that the case is well, but not perfectly, aligned with the calibration data. Conversely, a low-credibility singleton, as in Figure 6d, indicates an atypical or weakly supported prediction.

The performance difference between field strengths validated the method’s sensitivity to distribution shift: 3 T images consistently produced smaller prediction sets than 1.5 T images (baseline: 0.89 vs 0.94), reflecting the model’s greater confidence when operating within its training distribution. This behavior persisted across all degradation types, with relative responses to additional shift remaining nearly identical (83.4% increase for 1.5 T vs 79.9% for 3 T under maximum blur), suggesting robust scaling of uncertainty regardless of baseline distribution mismatch.

## 4 Discussion

Our results demonstrate that class-conditional CP offers a principled solution to a critical challenge in medical AI deployment: providing reliable UQ for minority classes representing less common but clinically important conditions. The significant failure of marginal CP for MS patients—with 100% severe undercoverage despite valid marginal guarantees—exemplifies how standard UQ methods can fail precisely where they are most needed. In clinical practice, where diseases require timely diagnosis and treatment decisions carry significant consequences, such systematic undercoverage of the minority class renders the AI system clinically unusable, regardless of its overall accuracy.

The success of class-conditional methods in mitigating this failure has direct implications for clinical deployment. By computing separate conformity scores for each diagnostic class, the approach ensures that UQ remains valid within each patient subpopulation. The observed trade-off—modestly increased undercoverage for healthy controls in exchange for dramatically improved coverage of MS patients—represents a clinically acceptable compromise. Healthy controls maintained coverage above 89% with zero severe undercoverage, while MS patients saw severe undercoverage frequency drop from near-universal to manageable levels (22-37% of runs).

The increased coverage variance for minority classes reveals a fundamental statistical constraint rather than a methodological limitation. Our theoretical analysis confirms that this variance follows directly from the Beta-binomial distribution governing finite-sample coverage, with variance inversely proportional to calibration set size. This finding has important practical implications: while class-conditional methods successfully prevent systematic bias against minority classes, they cannot overcome the inherent uncertainty of statistical inference from limited samples.

This constraint manifests differently across deployment contexts. The 1.5 T dataset, with only 7% MS prevalence, showed substantially higher coverage variance than the 3 T dataset with 14% prevalence. This dose-response relationship suggests that even modest improvements in minority class representation can yield significant gains in prediction reliability. For clinical deployment, this implies that targeted data collection focusing on underrepresented conditions may provide greater value than general dataset expansion.

The robust maintenance of coverage guarantees under distribution shift—from field strength differences to synthetic degradations—validates conformal prediction’s suitability for heterogeneous clinical environments. The method’s ability to appropriately scale uncertainty with distribution shift severity addresses a key deployment challenge: models trained at specialized academic centers must often operate in community settings with different equipment, protocols, and patient populations.

Crucially, the monotonic relationship between distribution shift and prediction set size offers an unexpected benefit: the prediction intervals themselves become diagnostic tools for data quality assessment. Clinicians can immediately recognize when image quality or acquisition parameters deviate from training conditions through expanded prediction sets, enabling appropriate adjustments to diagnostic confidence. This self-calibrating behavior aligns with radiological practice, where image quality directly influences diagnostic certainty.

The challenges we observe in MS diagnosis exemplify a fundamental characteristic of healthcare AI: severe class imbalance is the norm rather than the exception in clinical screening applications. Population-level disease prevalence often falls well below 5% for conditions requiring early detection, creating inherent tensions between statistical performance metrics and clinical utility. Whether screening for rare cancers, genetic disorders, early-stage neurodegenerative diseases, or acute cardiac events, AI systems must reliably identify the few positive cases among overwhelmingly healthy populations. This systematic imbalance renders standard machine learning approaches and uncertainty quantification methods inadequate, as they optimize for overall accuracy at the expense of minority class reliability. The class-conditional framework we demonstrate here thus addresses not merely a technical challenge but a pervasive barrier to clinical AI deployment across specialties. As healthcare systems increasingly adopt AI for population screening and risk stratification, methods that explicitly account for and mitigate effects of extreme class imbalance become essential infrastructure for equitable and effective care delivery.

While traditional methods for handling class imbalance such as synthetic oversampling (SMOTE), undersampling, or cost-sensitive learning can improve model training, they fundamentally differ from class-conditional CP in both requirements and objectives [48]. Resampling techniques require retraining models from scratch, making them incompatible with the black-box commercial AI systems already deployed in many clinical settings. Moreover, these approaches optimize point prediction accuracy rather than providing calibrated uncertainty estimates, a critical distinction when clinical decisions require explicit confidence assessment. Synthetic data generation may inadvertently introduce artifacts or unrealistic examples that compromise model generalization, while aggressive undersampling discards valuable majority class information. Perhaps most importantly, none of these training-time interventions provide the formal statistical guarantees that CP offers. Class-conditional CP uniquely combines post-hoc applicability, allowing deployment on existing models without modification, with mathematically rigorous coverage guarantees that hold even under distribution shift. Rather than competing approaches, we view training-time class balancing and deployment-time uncertainty quantification as complementary strategies: the former improving model discrimination, the latter ensuring reliable confidence assessment for clinical decision-making.

### 4.1 Limitations and Future Work

Several limitations constrain the immediate clinical deployment of these methods. First, while class-conditional approaches improve coverage balance, they require sufficient calibration data for each class—a challenge for ultra-rare conditions. Second, the substantial increase in prediction set size under severe distribution shift (>80% expansion) can render predictions clinically uninformative, even if statistically valid. Third, the current framework assumes a fixed deployment distribution, whereas clinical practice involves continuous evolution of imaging protocols and patient populations.

The interpretability gap between statistical guarantees and clinical decision-making remains significant. While CP provides mathematically rigorous uncertainty intervals, translating these into actionable clinical insights requires careful consideration of context. A prediction set containing both diagnostic options may be statistically valid but clinically ambiguous without additional risk stratification or decision support.

Our results suggest a path toward more reliable clinical AI deployment through hierarchical UQ. In resource constrained settings, primary screening could use marginal methods for efficiency, with class-conditional refinement for flagged cases. Prediction set size could trigger quality control protocols, prompting image reacquisition or specialist review when uncertainty exceeds clinical thresholds. Integration with electronic health records could provide contextual priors, potentially reducing prediction set sizes while maintaining coverage guarantees.

Future work should address several critical extensions. Adaptive conformal methods could handle evolving clinical distributions without full recalibration. Multi-dimensional extensions beyond binary classification would support differential diagnosis scenarios. Integration with active learning could prioritize acquisition of calibration data for underrepresented conditions. Most importantly, prospective clinical trials must validate whether these statistical guarantees translate into improved clinical outcomes.

### 4.2 Methodological Contributions and Broader Impact

This work contributes empirical evidence that class-conditional CP can address fundamental challenges in medical AI deployment: class imbalance, distribution shift, and the need for reliable UQ. By demonstrating these capabilities in a clinically relevant setting—MS diagnosis where early detection significantly impacts patient outcomes—we provide a template for applying these methods across medical domains characterized by rare disease detection and heterogeneous deployment conditions.

The open-source implementation accompanying this work enables researchers to apply and extend these methods to other medical imaging tasks. Our systematic evaluation framework, spanning coverage analysis, distribution shift assessment, and finite-sample characterization, provides a blueprint for rigorous UQ in clinical AI systems. As healthcare increasingly relies on AI-assisted diagnosis, such principled approaches to UQ become essential infrastructure for safe and effective deployment.

## 5 Conclusions

This study demonstrates that class-conditional CP provides a principled approach to UQ that addresses critical challenges in deploying AI systems for disease detection. Our systematic evaluation across 100 Monte Carlo experiments reveals both the promise and fundamental constraints of these methods when confronting the dual challenges of severe class imbalance and distribution shift characteristic of real-world clinical deployment.

The catastrophic failure of marginal CP for MS diagnosis—with 100% severe undercoverage at 1.5 T despite maintaining valid population-level guarantees—underscores why standard UQ approaches prove inadequate for clinical applications where clinical conditions carry disproportionate consequences. Class-conditional methods successfully mitigated this failure, reducing severe undercoverage frequency while maintaining coverage for the majority class. This improvement comes at the cost of increased coverage variance for minority classes, a fundamental statistical property we confirmed through theoretical analysis.

Our findings reveal that CP’s response to distribution shift—monotonically increasing prediction set sizes while maintaining coverage validity—offers an unexpected clinical utility. The 80% expansion in prediction set size under severe image degradation provides an implicit quality assessment mechanism, alerting clinicians when input data deviates substantially from training conditions. This self-calibrating behavior aligns naturally with clinical practice, where diagnostic confidence necessarily varies with image quality and acquisition parameters.

The work also exposes important limitations that must be addressed before widespread clinical deployment. The substantial coverage variance for minority classes, while theoretically expected, may prove challenging in ultra-rare conditions where calibration data remains scarce. The tension between statistical validity and clinical utility becomes apparent when prediction sets expand to include multiple diagnostic possibilities, providing mathematical rigor but limited actionable guidance. These challenges suggest that CP should complement rather than replace clinical judgment, serving as a principled framework for quantifying and communicating model uncertainty rather than a standalone decision-making tool.

Moving forward, several research directions warrant investigation. Adaptive conformal methods that accommodate evolving clinical distributions without full recalibration could address the practical constraints of deployment in dynamic healthcare environments. Extensions to multi-label and ordinal outcomes would better reflect the complexity of differential diagnosis and disease staging. Integration with active learning frameworks could guide strategic acquisition of calibration data for underrepresented conditions, potentially reducing the sample size requirements for reliable UQ.

This work contributes to the broader effort of developing trustworthy AI for healthcare by demonstrating that principled UQ methods can be successfully adapted to address the unique challenges of clinical deployment. These methods are directly applicable to clinical AI systems currently deployed or under development, requiring only post-hoc calibration without model retraining or architectural modifications. The open-source implementation and the evaluation framework we provide offer a foundation for researchers to extend these methods to other medical imaging applications where rare disease detection and distribution shift pose similar challenges. As the medical community increasingly relies on AI-assisted diagnosis, establishing rigorous approaches to UQ becomes not merely a technical refinement but an ethical imperative for safe and equitable healthcare delivery.

## CRediT Author Contribution Statement

**Alexander Millar:** Conceptualization, Data curation, Formal analysis, Funding acquisition, Investigation, Methodology, Project administration, Resources, Software, Supervision, Validation, Visualization, Writing – original draft, Writing – review & editing. **Cortnee Roman**: Conceptualization, Formal analysis, Investigation, Methodology, Resources, Supervision, Validation, Writing – review & editing. **Ramkiran Gouripeddi:** Formal analysis, Funding acquisition, Investigation, Methodology, Project administration, Supervision, Validation, Writing – review & editing. **Julio Facelli:** Conceptualization, Formal analysis, Funding acquisition, Investigation, Methodology, Project administration, Resources, Supervision, Validation, Writing – review & editing.

## Acknowledgments

This work was supported by the Department of Defense (DoD) SMART Scholarship-for-Service Program (AM) and the National Center for Advancing Translational Sciences of the National Institutes of Health under Award Number UM1TR004409 (JF, RG). The content is solely the responsibility of the authors and does not necessarily represent the official views of the Department of Defense or the National Institutes of Health.

## Data Availability

To foster transparency and reproducibility, this study relied exclusively on openly accessible brain MRI datasets. The IXI dataset (RRID:SCR_005839) is accessible at http://brain-development.org/ixi-dataset/ (direct download: <u>IXI-T2.tar</u>). The 2015 ISBI Longitudinal MS Lesion Segmentation Challenge dataset is available through the SmartStats website (https://smart-stats-tools.org/lesion-challenge) following free account registration. The Muslim et al. MS dataset is hosted on Mendeley Data (https://doi.org/10.17632/8bctsm8jz7.1). No new data were generated in this study.

## Code Availability

To support reproducibility and transparency, all code used in this study has been made available at https://github.com/illato/mricp. The repository includes all necessary scripts and documentation to reproduce the analyses presented in this article.

## Ethics Statement

Ethical approval and informed consent were not required for this study because it relied exclusively on secondary analysis of openly accessible, fully anonymized datasets (IXI, ISBI 2015, and the Muslim et al. cohorts). All original data collection procedures were conducted by the respective dataset creators in compliance with their local relevant laws and institutional guidelines. The privacy rights of human subjects were observed, and no new human subject data were generated in the course of this research.

## Declaration of Generative AI and AI-assisted Technologies in the Writing Process

During the preparation of this work the authors used ChatGPT and Claude to improve readability and concept presentation. After using these tools, the authors reviewed and edited the content as needed. The authors take full responsibility for the content of the published article.

## Supplementary Material

**Table S1.** Quantitative assessment of conformal prediction performance under distribution shift.

| Field Strength† | Variant | Mean PS Size | PS Size Change‡ | Mean Coverage | Coverage Deviation§ |
| --- | --- | --- | --- | --- | --- |
| **1.5 T MRI** | Baseline | **0.94 ± 0.04** | — | 89.8% ± 3.8% | -0.2% |
|  | Blur SD1 | 0.94 ± 0.04 | +0.0% | 89.6% ± 3.8% | -0.4% |
|  | Blur SD2 | 0.99 ± 0.07 | +4.9% | 89.5% ± 3.6% | -0.5% |
|  | Blur SD3 | 1.35 ± 0.10 | +43.9% | 89.8% ± 3.2% | -0.2% |
|  | Blur SD4 | 1.72 ± 0.06 | **+83.4%** | 89.8% ± 2.6% | -0.2% |
|  | Contrast (AHE) | 0.98 ± 0.07 | +4.7% | 89.7% ± 3.8% | -0.3% |
|  | Contrast (HE) | 1.72 ± 0.06 | +82.8% | 89.3% ± 3.1% | -0.7% |
|  | Contrast (IA) | 0.95 ± 0.05 | +1.2% | 89.6% ± 3.8% | -0.4% |
| **3 T MRI** | Baseline | **0.89 ± 0.04** | — | 88.5% ± 3.8% | -1.5% |
|  | Blur SD1 | 0.90 ± 0.04 | +0.8% | 88.7% ± 3.6% | -1.3% |
|  | Blur SD2 | 0.94 ± 0.04 | +4.8% | 89.0% ± 3.3% | -1.0% |
|  | Blur SD3 | 1.18 ± 0.05 | +32.1% | 89.3% ± 2.8% | -0.7% |
|  | Blur SD4 | 1.61 ± 0.06 | **+79.9%** | 89.8% ± 2.7% | -0.2% |
|  | Contrast (AHE) | 0.90 ± 0.03 | +1.2% | 89.6% ± 2.9% | -0.4% |
|  | Contrast (HE) | 1.71 ± 0.03 | +91.1% | 89.8% ± 2.9% | -0.2% |
|  | Contrast (IA) | 0.91 ± 0.04 | +1.7% | 89.3% ± 3.3% | -0.7% |

†Model trained on 3 T baseline data; 1.5 T represents inherent distribution shift. ‡Relative change from baseline. §Deviation from 90% target. AHE: Adaptive histogram equalization; HE: Histogram equalization; IA: Intensity adjustment; PS: Prediction set.

